# Detecting COVID-19 Related Pneumonia on CT Scans using Hyperdimensional Computing

**DOI:** 10.1101/2021.05.21.21257631

**Authors:** Neftali Watkinson, Tony Givargis, Victor Joe, Alexandru Nicolau, Alexander Veidenbaum

**Affiliations:** The Donald Bren School of Information and Computer Sciences, University of California, Irvine Donald Bren Hall, 6210, Irvine, CA 92697; The Regional Burn Center, UCI Medical Center, 101 The City Dr S, Orange, CA 92868

## Abstract

Pneumonia is a common complication associated with COVID-19 infections. Unlike common versions of pneumonia spread quickly through large lung regions, COVID-19 related pneumonia starts in small localized pockets before spreading over the course of several days. This makes the infection more resilient and with a high probability of developing acute respiratory distress syndrome. Because of the peculiar spread pattern, the use of pulmonary computerized tomography (CT) scans was key in identifying COVID-19 infections. Identifying uncommon pulmonary diseases could be a strong line of defense in early detection of new respiratory infection-causing viruses. In this paper we describe a classification algorithm based on hyperdimensional computing for the detection of COVID-19 pneumonia in CT scans. We test our algorithm using three different datasets. The highest reported accuracy is 95.2% with an F1 score of 0.90, and all three models had a precision of 1 (0 false positives).

## 1. Introduction

On January 2020, the World Health Organization declared a global emergency due to a novel coronavirus that had started in the regions of Wuhan, China and rapidly spread around the world. Symptoms include fever, headache, loss of smell, difficulty breathing, among others [1]. In the early stages of creating a diagnosis protocol, medical experts were specifically interested in the effects SARS-CoV-2 (COVID-19) had in the patient’s lungs. In severe cases, lungs get inflamed, filled with fluid and debris, causing what is known as pneumonia [2]. Before any quick test was developed to identify the infection, hospitals relied on computerized tomography (CT) scans [3] and using lower respiratory tract samples [4]. However, accuracy among expert radiologist on differentiating COVID-related pneumonia from typical pneumonia can vary from 97% down to 67% [5].

At the time of writing, various blood and saliva tests have been developed, but their accuracy has been the focus of debate. While most tests seem to emphasize sensitivity over specificity [6], CT scans remain the most accurate way to confirm symptomatic infections, especially when dealing with in-hospital settings and with patients with preexisting lung related conditions [7].

Recent research has focused on applying artificial intelligence to the challenge of detecting COVID-19 in CT scans [8]. The work of Soares, et al. [9] aims to build an explainable deep learning network using CT scans collected from patients across several hospitals in Sao Paulo, Brazil. They released their dataset containing images from healthy patients, patients with COVID-19 related pneumonia, and patients with other pulmonary issues. Similarly, He, et al. [10] released a dataset containing ct-scans of healthy patients and patients diagnosed with COVID-19 that were data mined from other studies. Additionally, Rahimzadeh, et. al. [11] released a dataset of CT scan sequences from the same source and introduces a deep learning based model to identify COVID-19 infections.

In this paper we describe a hyperdimensional (HD) computing [12] approach for identifying CT scan images as suspicious of COVID-19 using image classification. We train and test the models using three separate datasets. Our approach achieves up to 95% classification accuracy and report 0 false positives. We compared our models with those originally published along the datasets and present a thorough discussion on the advantages and disadvantages of our approach.

The rest of the paper is organized as follows:

- **Methodology** explains our HD computing implementation and justifies decisions made during the design of the classification model.
- **Results** describes the resulting accuracy with each of the CT scans datasets and compares them with the models described in the original papers that published the datasets.
- In **Discussion** we justify our approach and talk about the future work.
- **Conclusion** summarizes our findings and future work.

## II. Methodology

Our algorithm relies on HD computing to encode one gray scale image sample per CT scan into a *hypervector*, which for practical purposes it is a very long vector. For this work we use vectors with 10 thousand elements. Once encoded we can measure the distance between hypervectors. The predicted class for unknown encoded images will be that of the closest known hypervector. In this section we describe the specific characteristics of our implementation.

### A. HD Computing for Image Classification

This is not the first time HD computing is used for image classification [13]–[15]. There different ways to implement it, but we follow an algorithm very similar to the one described by Yang, et al. [16]. We use bipolar hypervectors (every element has a value of 1 or -1) and encode both the intensity and the position of every pixel. After obtaining both hypervectors, we bind them through multiplication. Then we combine all the pixel hypervectors through majority voting [12] to generate a single hypervector for the image.

### B. Encoding

For creating the hypervectors, we use orthogonal or un-correlated encoding [13]–[15] to represent the position of each pixel and linear or correlated encoding [17] to represent the pixel intensity. This approach requires that the images have the same number of pixels (equal in size) and a similar contrast ratio in order to reduce noise.

#### 1) Preprocessing

Two of the three datasets [9], [10] have images with various sizes. We resize those images to be 200 pixels high and 300 pixels wide. The third dataset [11] contains images from the same source and are all sized the same (512 pixels high and wide). We used OpenCV’s [18] Python library for resizing and for normalizing their histogram in order to get a similar contrast ratio across the datasets. Preprocessing and image filtering is not unusual in machine learning-based image processing tasks [19]–[21], and commonly used for face [22] and object [23] detection. During normalization, we downscale the intensity values of the third dataset from the original 16-bit values down to 8-bit. Therefore, for all three datasets, each pixel has a value in the range of 0-255.

#### 2) Training

Once all the images are preprocessed, and the dimensionality (10k) and type (bipolar) of the hypervectors has been set, the training phase proceeds as follows:

1. Identify the magnitude of features. Since all the images are the same size after processing, then we have 300 by 200 pixels or 60,000 features for datasets 1 [9] and 2 [10], and 512 by 512 (262,144) pixels for dataset 3 [11].
2. For each pixel, encode the position and the intensity hypervector and bind them.
3. Combine all the pixel hypervectors of a single image using majority voting.
4. Add all the feature vectors using majority voting.
5. Store the image hypervector keeping record of what class (healthy or COVID-19) it belongs to.

#### 3) Testing

We separate a subset of the images that are not part of the training phase. This subset is called the testing subset and is used to evaluate the model. For this work we use a 70-30 split, which means that 70% of the images will be used for training and the remaining 30% will be used for testing. They all go through the same encoding process with the only difference that the testing images’ class is set to that of the closest training hypervector. We then compare this class with the actual class of the original image and derive the accuracy of the model in correctly predicting the testing images. This type of validation is just one of the widely documented [24] techniques for model validation.

## III. Results

Imaging data from CT scans are generally stored using the DICOM formatting [25] that contains information such as patient data. For the three datasets, the images have been scrubbed of identifying data and extracted as Portable Network Graphics (PNG) images. Axial CT scans are done from the perspective of the axial or transverse plane, along or perpendicular to the median plane. In other words, with the patient lying on their back, slices are collected starting from the upper lobe of the lungs (closest to the patient’s head) towards the lower lobe (closest to the patient’s waist). Figure 1 shows an image sample for a single patient. Ranges and slice sizes vary for each patient. We chose to focus on the mid section with the right major fissure in focus and the trachea splitting into the main bronchi (b) since this was fairly consistent among all patients for all datasets.

**Fig. 1.**
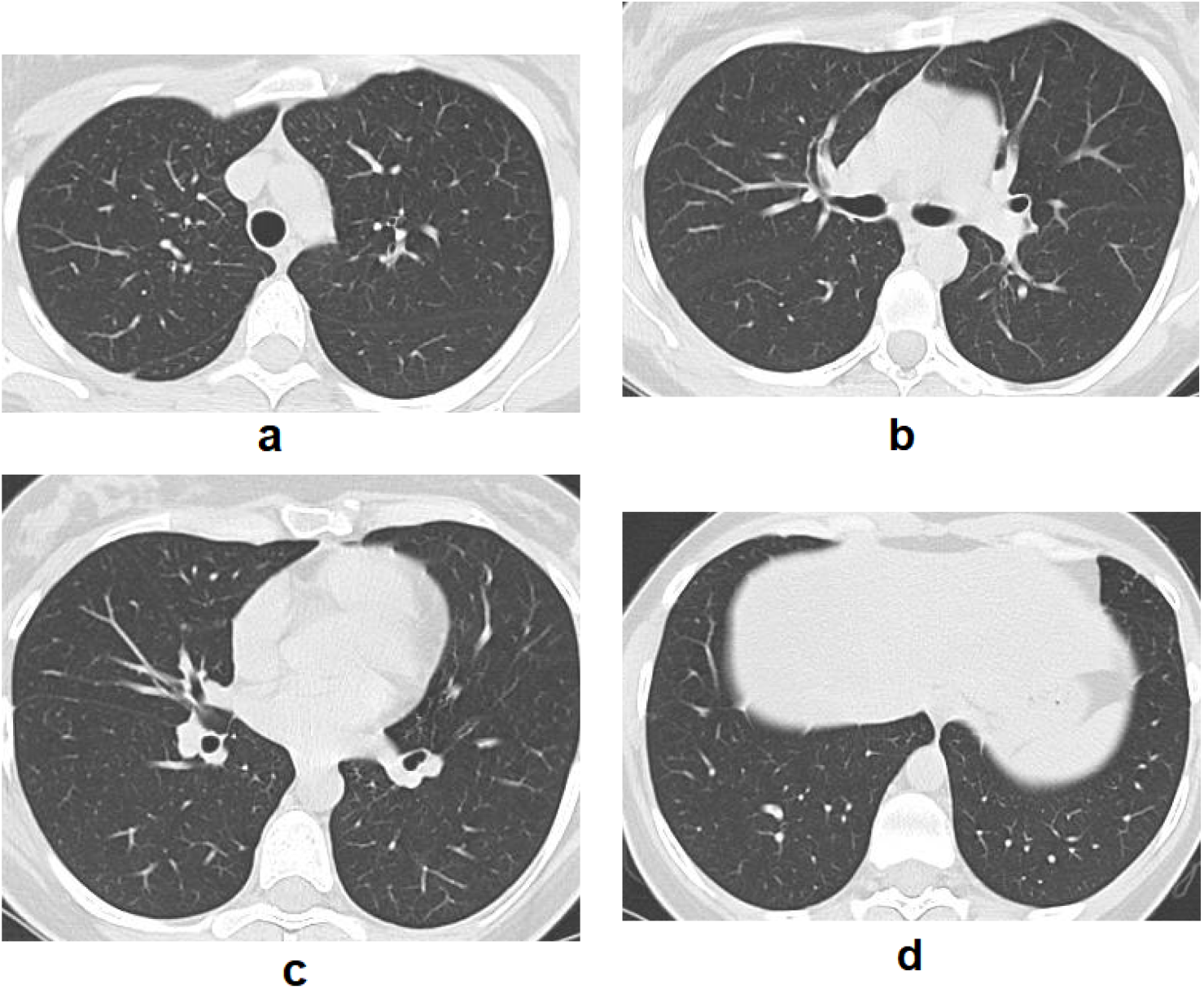
CT scan image sample from one patient showing the upper lobe and the trachea in the middle of the image(a), the mid section of the lung showing the anterior segment in focus and the trachea splitting into the main bronchi (b), the lower lobe section with inferior lobar bronchi (c), and the basal segments of the lower lobe with the diaphragm starting to appear (d)

We built three models, one for each dataset. Due to qualitative issues (some images having very poor quality), for dataset 1 [9] we handpicked one slice per patient that met a minimum qualitative standard (lungs clearly visible and image size no smaller than 150 by 150 pixels). Through this process we obtained 80 images from patients with COVID-19 and 46 images from healthy patients. From dataset 2 [10], we sued all the images which totalled 350 with COVID-19 and 398 without. From dataset 3, we used one image per patient from all 96 patients with COVID-19 and randomly sampled 109 healthy patients in order to keep the dataset balanced. The data set has 46 scans from healthy patients, 80 scans from patients diagnosed with COVID-19, and 67 from patients with non-COVID-19 pulmonary illnesses. Table I shows the population distribution for each dataset and Figures 2 and 3 show an image sample of a healthy and a COVID-19 CT scan image respectively.

**TABLE I.**
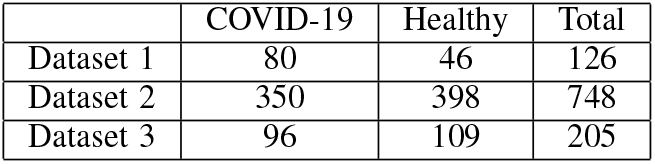
This table shows the population distribution for each of the datasets

|  | COVID-19 | Healthy | Total |
| --- | --- | --- | --- |
| Dataset 1 | 80 | 46 | 126 |
| Dataset 2 | 350 | 398 | 748 |
| Dataset 3 | 96 | 109 | 205 |

**TABLE II.**
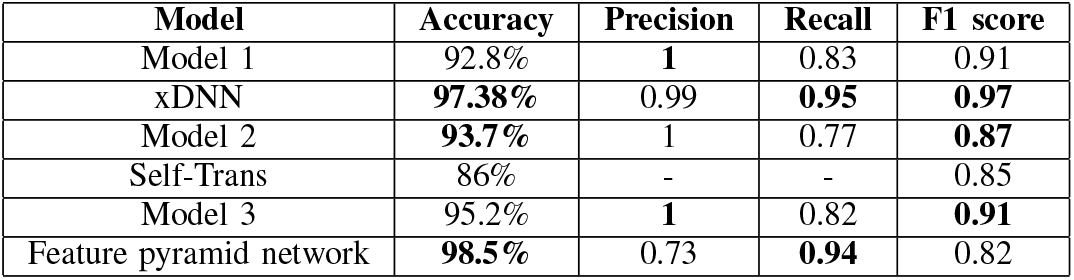
Accuracy, precision, recall and f1 score for our models compared to the model used for each of the published datasets in their original papers. Model 1 was used for dataset 1, model 2 for dataset 2 and model 3 for dataset 3.

| Model | Accuracy | Precision | Recall | F1 score |
| --- | --- | --- | --- | --- |
| Model 1 | 92.8% | **1** | 0.83 | 0.91 |
| xDNN | **97.38%** | 0.99 | **0.95** | **0.97** |
| Model 2 | **93.7%** | 1 | 0.77 | **0.87** |
| Self-Trans | 86% | - | - | 0.85 |
| Model 3 | 95.2% | **1** | 0.82 | **0.91** |
| Feature pyramid network | **98.5%** | 0.73 | **0.94** | 0.82 |

**Fig. 2.**
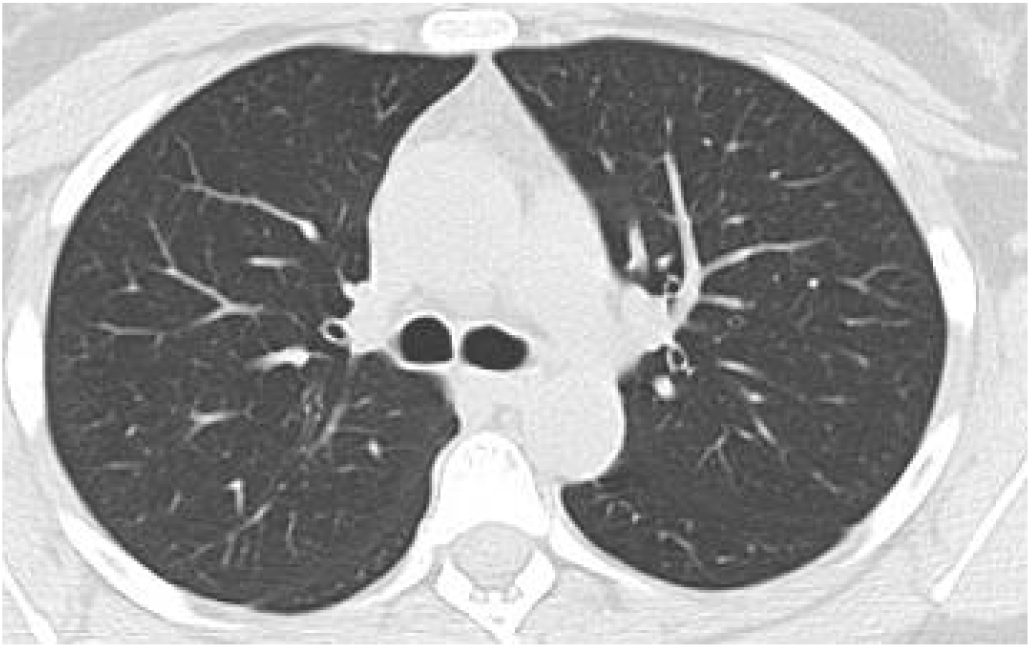
Image from a CT scan of a patient with healthy lungs showing minimal haziness and clear definition of the arteries

**Fig. 3.**
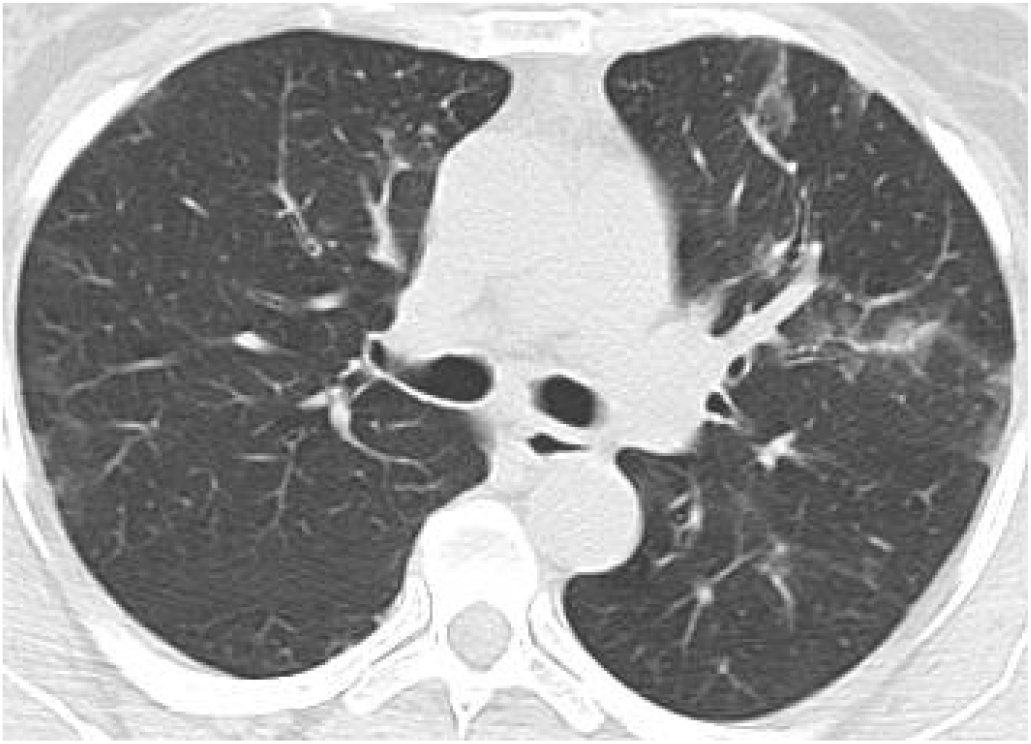
Image from a CT scan of a patient with COVID-19. The haziness is indicative of pneumonia and the pattern is consistent with that of patients with a COVID-19 infection.

### A. Comparing to expert radiologists

For the models corresponding to each dataset we derived the classification accuracy, the precision, recall and F1 score and compared them to the models used in the original papers for each dataset. These results are in Table **??**. It is important to note that the model originally presented in the work of Soares, et al. [9] has a data leakage issue where images where randomly split without separating them per patient. This means that images from the same patient can appear in the training and the validation set. All of our models had a precision of 1 which means that there were no false positives.

## IV. Comparing to expert radiologists

The performance of the binary classification model is comparable to the median accuracy values presented in [5]. However, there are key differences between the two studies. In [5], CT scans from COVID-19 patients with no abnormalities were discarded. Additionally, radiologist had access to the full scan. For the model presented in this chapter, only one slice is being analyzed and all COVID-19 are included, without discarding non-anomalous images. [11] observed in a separate experiment that radiologists where only 70% accurate in detecting COVID-19 infections in CT scans. Future research needs to be done to discover an encoding that is not dependent upon pixel position and that could be implemented to three-dimensional images.

## V. Discussion

HD computing has proven to be an efficient machine learning approach for many domains, specially those that are data constrained [13], [17], [26], [27]. In this work we show that classification of CT scan images is another use for it. Our models consistently achieve over 92% classification accuracy and surpasses deep learning based models in at least one of the metrics. An advantage that HD computing has that we don’t discuss in this paper is that it is computationally efficient [12]. Additionally we are reducing the input dimensionality from up to 262 thousand pixels (for the images that are 512 by 512 pixels) down to 10 thousand bipolar digits.

On the other hand, our HD computing models perform well with little to medium amounts of data. Dataset 1 only contains 126 images and dataset 2 contains 748. This means that this approach has both the potential of being used to detect new unseen anomalies where data is scarce, or scale and improve as more data is made available. However, there is still much work to do in order to properly assess this, in addition to incorporating a full CT scan sequence as a hypervector, with the option of adding the patient’s data as well.

## VI. Conclusion

In this paper we describe a hyperdimensional computing image classification approach to classifying images from pulmonary CT scans across two classes: Healthy and with COVID-19 related pneumonia. We test our approach through three models, each one applied to a different dataset. All the models achieve over 92% classification accuracy and beat the state of the art models that were used originally on these datasets by at least one metric (accuracy, precision, recall and F1 score). Additionally, none of the models generate any false negatives and greatly reduce the input dimensionality. We argue that this approach has the potential of detecting new pulmonary diseases but also scales well when more data is made available.

## Data Availability

The reference to the data sources is included in the paper

